# Demographic and Clinical Characteristics of Pediatric COVID-19 in Arkansas: March-December 2020

**DOI:** 10.1101/2022.06.18.22276587

**Authors:** Sara C. Sanders, Maxwell D. Taylor, Jacob Filipek, Dustin Williford, Cindy Nguyen, Charalene R. Fisher, Stephanie M. Scheffler, Emily S. Smith, Phoebe Martin, Rebecca L. Latch, Jessica Snowden, Chang L. Wu, Rebecca M. Cantu

## Abstract

The COVID-19 pandemic reached the United States in early 2020 and spread rapidly across the country. This retrospective study describes the demographic and clinical characteristics of 308 children presenting to an Arkansas Children’s emergency department or admitted to an Arkansas Children’s hospital with COVID-19 in the first ten months of the COVID-19 pandemic, prior to the emergence of clinically significant variants and available vaccinations. Adolescents aged 13 and older represented the largest proportion of this population. The most common presenting symptoms were fever, gastrointestinal symptoms, and upper respiratory symptoms. Patients with multisystem inflammatory syndrome in children (MIS-C) had a longer length of stay than patients with acute COVID-19. Children from urban zip codes had lower odds of admission but were more likely to be readmitted after discharge. Nearly twenty percent of the study population incidentally tested positive for COVID-19. Despite lower mortality in children with COVID than in adults, morbidity and resource utilization are significant. With many Arkansas children living in rural areas and therefore far from pediatric hospitals, community hospitals should be prepared to evaluate children presenting with COVID-19 and to determine which children warrant transport to pediatric-specific facilities.

## Introduction

COVID-19, the disease caused by the novel coronavirus SARS-CoV-2, emerged in Asia in December 2019 and spread rapidly, leading to a global pandemic^1^. With wide variability in infection rates, symptoms, and illness severity, COVID-19 impacted various geographical regions across the world in different ways. The first confirmed COVID-19 case in Arkansas was identified in March 2020, with the first death in the state attributed to COVID-19 occurring 13 days later. In an effort to contain viral spread, various methods were employed including restricting the size of gatherings, closing casinos temporarily and public schools for the remainder of the school year, and limiting the availability of indoor services ^2^. The first pediatric admissions in Arkansas occurred a month after the first adult cases of COVID-19.

The pediatric population, those of 21 years of age and under, comprises approximately 26% of the U.S. population ^3^. Published data generally shows that COVID-19 severity, morbidity, and mortality is lower in the pediatric population than in older demographics ^1, 4^. Even so, 121 pediatric deaths in the United States were reported to the Centers for Disease Control and Prevention (CDC) in the first five months of the pandemic. Among children, those aged 10-20 years old had increased morbidity and mortality, as did children with at least one underlying medical condition ^3^.

Pediatric COVID-19 can present with a range of symptoms including fever, respiratory symptoms, and gastrointestinal symptoms, and many children have asymptomatic infection ^1, 5^. Additionally, children are at risk of post-COVID-19 complications, including multisystem inflammatory syndrome in children (MIS-C), myocarditis, and persistent symptoms sometimes referred to as “long COVID” ^6^. There are also racial and ethnic disparities in MIS-C even after controlling for COVID-19 and geographic variations ^7^. This study aims to describe the demographic and clinical characteristics of children with COVID-19 presenting to our pediatric emergency department or admitted to our pediatric hospitals in the first ten months of the COVID-19 pandemic.

## Methods

This is a retrospective chart review study of children diagnosed with COVID-19 who presented for care in a pediatric health system in Arkansas from March 1, 2020-December 31, 2020. The two-hospital health system consists of a main 336-bed freestanding children’s hospital with a 38-bed pediatric emergency department (ED) and a separate 24-bed community children’s hospital with a 24-bed ED in another portion of the state. The community hospital does not have an intensive care unit (ICU); the main children’s hospital has an ICU and has a transport service utilizing ground, helicopter, and fixed-wing air transport methods. Children with severe or escalating illnesses, or those requiring subspecialty care not available at the community hospital, are often transported to the main hospital approximately 200 miles away. As part of the same system, both hospitals utilize the same electronic health record, Epic (Epic Systems Corporation). Patients from both hospitals were included in the study. Patients who presented to the community hospital and then transported to the main hospital were counted as 1 visit. COVID-19 testing for children evaluated in the ED or admitted to the hospital consisted of Emergency Use Authorization (EUA)-approved nucleic acid amplification tests (NAAT), including Lyra SARS-CoV-2 (Quidel, San Diego, CA), Lyra Direct SARS-CoV-2 (Quidel, San Diego, CA), Aptima SARS-CoV-2 (Hologic, Bedford, MA), or Xpert Xpress SARS-CoV-2 (Cepheid, Sunnyvale, CA). The limit of detection for SARS-CoV-2 was 750 cp/mL (Lyra and Lyra Direct), 350 cp/mL (Aptima) and 250 cp/mL (Xpert Xpress). All NAAT was performed according to the manufacturer’s instructions. Per hospital policy, all admitted children underwent NAAT unless they had a documented NAAT from a referring hospital the same day.

Eligible patients under 21 years of age were identified in the EHR by an automatic report based on results of COVID NAAT and cross-referenced for accuracy with an electronic report of positive COVID-19 tests maintained by the hospital system’s Infection Prevention department. Manual chart review was performed to extract demographic and clinical characteristics. Rurality was estimated using rural-urban commuting area (RUCA) codes, dichotomized a priori as urban and non-urban ^8, 9^. Descriptive and logistic statistical analysis was performed utilizing SAS 9.4 (SAS Institute, Cary, NC). General linear modeling was performed with R (R Core Team (2021). R: A language and environment for statistical computing. R Foundation for Statistical Computing, Vienna, Austria. URL https://www.R-project.org/.). This study was approved as exempt by the University of Arkansas for Medical Sciences Institutional Review Board.

## Results

A total of 308 patients were identified for inclusion during the study period. The median age was 8.68 years (range 1 day - 20 years, interquartile range [IQR]: 0.84-15.74 years). The most common age group seen were adolescents aged 13 and older, comprising 37.7% of the study population, and the least frequently seen group were children ages 2-5, with 10.7% of the study population. Sixty-nine (22.4%) of the patients were from non-urban areas based on their home zip codes. Demographic characteristics including age, ethnicity, and sex did not significantly differ between admitted and non-admitted patients (Table 1). In our EMR, race was defined as black, white, and other, and in this division was significantly different between admitted and non-admitted patients (P<0.008); when reported as a binomial variable (white vs. non-white), this difference was no longer statistically significant. While many patients listed as “other” race were of Hispanic ethnicity, ethnicity was not significantly associated with admission. One patient in the 2-5-year age group died due to drowning, with COVID-19 as a presumed incidental diagnosis at the time of admission.

**Table 1.**
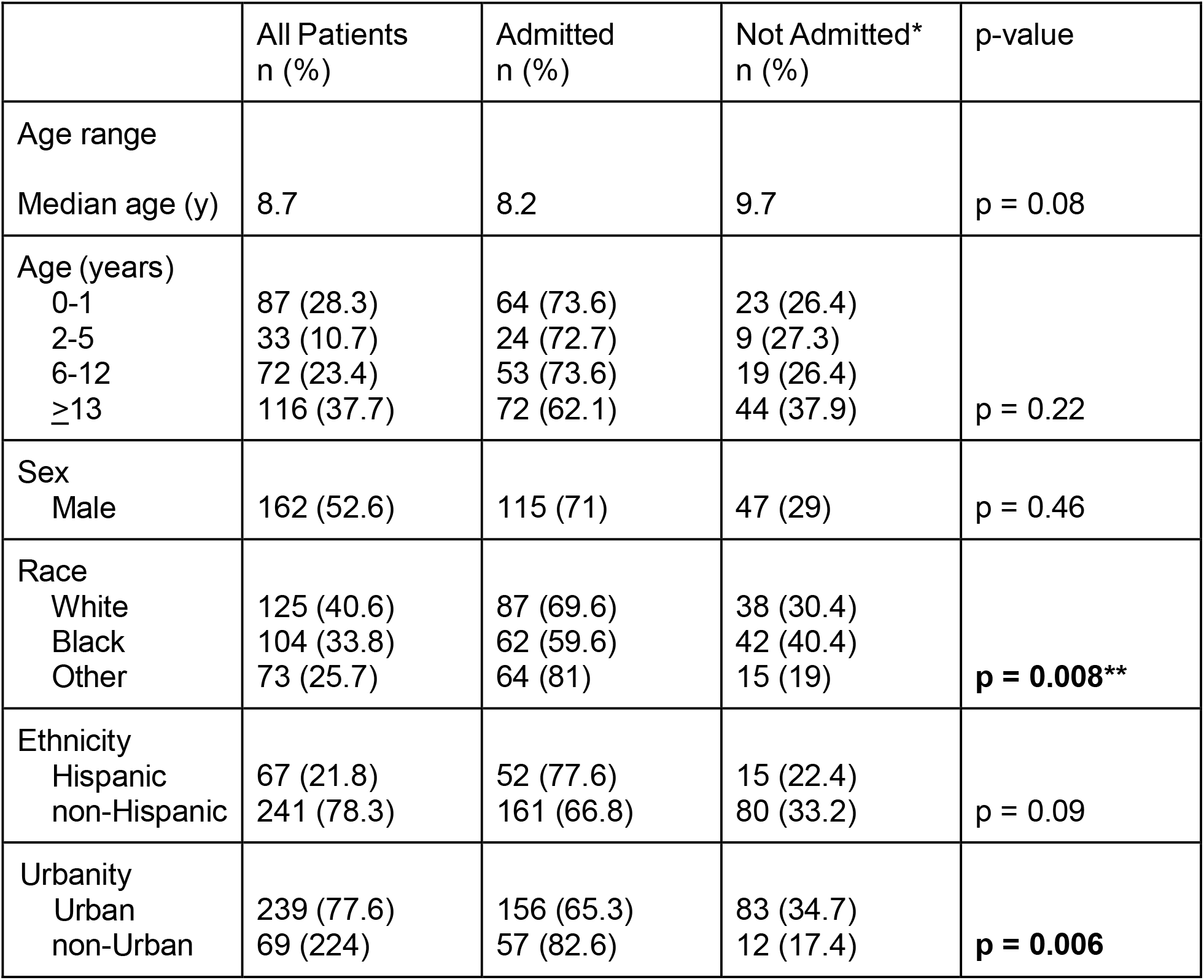
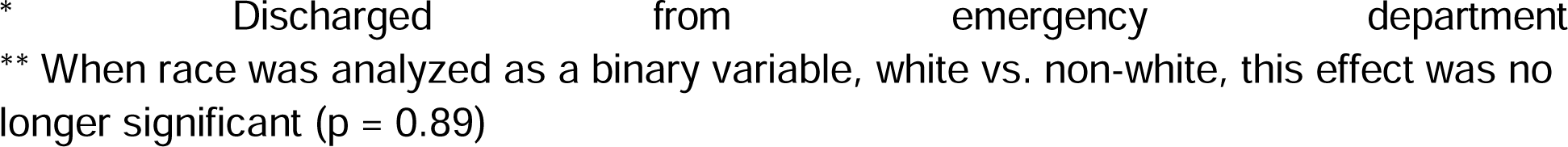
Demographic Characteristics and Likelihood of Admission and Readmission.

|  | All Patients<br>n (%) | Admitted<br>n (%) | Not Admitted*<br>n (%) | p-value |
| --- | --- | --- | --- | --- |
| Age range |  |  |  |  |
| Median age (y) | 8.7 | 8.2 | 9.7 | $p = 0.08$ |
| Age (years) |  |  |  |  |
| 0-1 | 87 (28.3) | 64 (73.6) | 23 (26.4) | $p = 0.22$ |
| 2-5 | 33 (10.7) | 24 (72.7) | 9 (27.3) |  |
| 6-12 | 72 (23.4) | 53 (73.6) | 19 (26.4) |  |
| ≥13 | 116 (37.7) | 72 (62.1) | 44 (37.9) |  |
| Sex |  |  |  |  |
| Male | 162 (52.6) | 115 (71) | 47 (29) | $p = 0.46$ |
| Race |  |  |  |  |
| White | 125 (40.6) | 87 (69.6) | 38 (30.4) | **$p = 0.008^{**}$** |
| Black | 104 (33.8) | 62 (59.6) | 42 (40.4) |  |
| Other | 73 (25.7) | 64 (81) | 15 (19) |  |
| Ethnicity |  |  |  |  |
| Hispanic | 67 (21.8) | 52 (77.6) | 15 (22.4) | $p = 0.09$ |
| non-Hispanic | 241 (78.3) | 161 (66.8) | 80 (33.2) |  |
| Urbanity |  |  |  |  |
| Urban | 239 (77.6) | 156 (65.3) | 83 (34.7) | **$p = 0.006$** |
| non-Urban | 69 (22.4) | 57 (82.6) | 12 (17.4) |  |

\* Discharged from emergency department
\*\* When race was analyzed as a binary variable, white vs. non-white, this effect was no longer significant ( $p = 0.89$ )

The average length of stay (LOS) was 6.5 hours in the ED-only subset (range 0.65-117 hours) of children evaluated in the ED but not admitted (31% of total study group; 95/308). Sixty-nine percent of patients (213/308) were admitted to the hospital, and 22% of admitted patients required ICU care. The mean LOS in patients admitted to the hospital was 109.5 hours (range 4.35-4205.9 hours). The median LOS of patients with an ICU stay was 157.27 hours (IQR: 6.59-15.75). Two admitted patients had outlier lengths of stay: (1) patient admitted to the burn unit for 61 days and incidentally found to be COVID-positive on admission; and (2) patient admitted due to premature birth to the neonatal intensive care unit for 175 days who became COVID-positive 2 months prior to discharge. A subanalysis of LOS for admitted patients was performed excluding these two outliers and yielded a mean LOS of 83.5 hours.

The most common presenting symptoms were fever (39.6%), gastrointestinal symptoms (27.6%), and upper respiratory symptoms (26%). Admission was significantly more likely in patients with presenting symptoms of lower respiratory symptoms including dyspnea and hypoxia (p<0.05), fever (p<0.001), or an incidental positive test unrelated to presenting symptoms (p<0.001) (Table 2). Of the pre-existing conditions evaluated, only asthma significantly altered risk of admission (OR 0.36, p<0.01). Nearly 20% of all patients in the study presented for admission or symptoms unrelated to COVID-19 and were incidentally found to be positive. Only 36% of patients were documented to have a known exposure to someone with COVID-19.

**Table 2.** Presenting Symptom and Likelihood of Hospital Admission.

|  | Number (%) | Odds Ratio (OR) |
| --- | --- | --- |
| Fever | 122 (39.6) | 3.55 (**$p<0.001$**) |
| Gastrointestinal | 85 (27.6) | 1.38 ($p=0.3$) |
| Upper respiratory | 80 (26) | 0.47 (**$p=0.015$**) |
| Lower respiratory | 62 (20.1) | 2.28 (**$p=0.02$**) |
| Incidental/Unrelated | 61 (19.8) | 8.83 (**$p<0.001$**) |
| Headache | 32 (10.4) | 0.62 ($p=0.25$) |
| Rash | 9 (2.9) | 0.79 ($p=0.76$) |
| Cardiac/Shock | 2 (0.65) | 0.97 ($p=0.98$) |

\* Patients could have more than one presenting complaint documented; numbers do not total 100%

MIS-C was diagnosed in 18 patients (5.8% of the total study population, 8.5% of admitted patients) with a median age of 8.59 years (range 2-18.6 years, IQR: 6.99-9.65 years). All patients with MIS-C were admitted to the hospital with 15 (83%) requiring ICU admission. The average LOS for MIS-C patients was 215 hours (range 112.5-344.8 hours). There were no mortalities in the MIS-C subset (Table 3).

**Table 3.** Patients With Multisystem Inflammatory Syndrome in Children (MIS-C)

|  |  |
| --- | --- |
| Age | Median 8.59 y (range 2-18.6 y, interquartile range: 6.99-9.65 y) |
| Race | White 7 (39%)<br>Black 8 (44%)<br>Other 3 (17%) |
| Ethnicity | non-Hispanic 15 (83%)<br>Hispanic 3 (17%) |
| Documented COVID-19 exposure | Yes 7 (39%)<br>No 11 (61%) |
| Medications | Intravenous immunoglobulin 16<br>Systemic corticosteroid 15 |
|  | Enoxaparin 15<br>Antibiotics 13<br>Pressors 11 |
| Presenting Symptom(s)* | Gastrointestinal 15<br>Fever 14<br>Headache 5<br>Rash 4<br>Upper respiratory (cough, congestion) 2<br>Dyspnea, hypoxia 1 |

\* Patients could have more than one presenting complaint documented

In patients with a positive COVID-19 NAA, the all-cause readmission rate was 7.1% in the first 7 days after discharge, and 15.6% over 30 days. The all-cause readmission rate for the main hospital during the study period was 5% at 7 days and 12% at 30 days. In multivariate regression analysis, no age, racial, or ethnic group was significantly more likely to require hospital admission or be readmitted after discharge (all p>0.05). Patients with urban zip codes were significantly less likely to be admitted (OR 0.4, 95% CI 0.2-0.78, p=0.006), but were significantly more likely to be readmitted within 7 days of discharge (p=0.035). There were no significant demographic predictors for 30-day readmission (all p>0.05).

## Discussion

This study describes demographic and clinical characteristics of pediatric patients at Arkansas Children’s Hospital during the first 10 months of the COVID-19 pandemic. All pediatric age ranges were represented, with the smallest group being children aged 2-5 years.

Our study identified a few unexpected findings. In our study population, 22.4% of children were considered non-urban based on RUCA codes. Children from non-urban areas were significantly more likely to be admitted than children with urban zip codes. However, children with urban zip codes had significantly higher 7-day readmission rates. This may indicate the effect of locality and ease of accessing medical care in the disposition planning process and determining need for hospital admission and readiness for discharge. Literature on adult patients suggests that rural areas have proportionally higher incidence rates of COVID-19 than urban areas, with populations with high rates of risk factors for severe COVID-19 infections ^10^.

Our population had an all-cause 7-day readmission rate of 7.1%. For comparison, Auger et al ^11^ estimated all-cause pediatric readmission rate of 5.1% in 2019. Contrary to what may be expected, children with asthma had less risk for admission. This may relate to limitations of documentation based on ICD-10 coding and problem lists rather than a true clinical difference.

Previous literature has reported incidence of MIS-C of 316 cases per 1 million pediatric COVID-19 cases ^12^. In our study, MIS-C was diagnosed in 8.5% percent of admitted patients, with 83% requiring ICU care at some point during their stay. Patients with MIS-C had a 157% longer mean LOS than admitted patients without a diagnosis of MIS-C (excluding the 2 outlier LOS data points as described in Results). The incidence of MIS-C and its impact may change with the emergence of the Delta, Omicron, or other variants and the availability of COVID-19 vaccines in different age groups. These findings underscore that while mortality may not be high in children with COVID, morbidity and resource utilization are significant.

Overall, outcomes in the study population were good. There was 1 death not seemingly related to COVID-19 (drowning, with incidental COVID-19), with the remainder of patients surviving to discharge. Follow-up data was not captured in this study. All-cause 7- and 30-day readmission rates were higher than baseline readmission rates for the study period. Potential causes for increased readmission rates after COVID are an area for future research.

Interestingly, only 36% of patients had a documented known COVID-19 exposure. Nearly 20% of the study population presented for symptoms or problems unrelated to COVID. These findings emphasize both the relatively mild presentation in the pediatric population and the importance of a low threshold to test for COVID-19 in the presence of exposure or any symptoms ^13^, as asymptomatic or mildly symptomatic children can also contribute to household transmission of COVID-19 ^14^.

Strengths of this study include the granularity of data available through manual chart review. Due to the large catchment area of this healthcare system, this study likely included most eligible pediatric patients who required hospitalization. Additionally, rural states have been disproportionately impacted by the COVID-19 pandemic and reflect admission/readmission patterns that may not be apparent in more urban states.

As a retrospective chart review, this study is limited by its reliance on appropriate documentation and coding. This study included the first 10 months of the pandemic so the time period does not include the emergence of the Delta variant of COVID, which has been shown to impact the pediatric population more than the initial strain. Additionally, vaccination against COVID-19 was not available for any pediatric age group during the study period. Future research into the impact of SARS-CoV-2 variants and vaccination of children and adolescents is warranted.

## Conclusion

Although less common than in adults, COVID-19 significantly impacts the children of Arkansas. We identified no specific demographic or clinical variable to predict the likelihood of hospital admission, emphasizing the importance of considering COVID-19 as a risk for all children. Further research is needed on the impact of new COVID-19 variants and the availability of COVID-19 vaccines. With many Arkansas children living in rural areas and therefore far from pediatric hospitals, community hospitals should be prepared to evaluate children presenting with COVID-19 and to determine which children warrant transport to pediatric-specific facilities. This study found that children typically had a mild presentation with a low rate of known exposure, supporting a low clinical index of suspicion and a high testing rate given the large number of children who are unknowingly exposed and infected.

## Data Availability

All data produced in the present study are available upon reasonable request to the authors.

## Acknowledgements

The authors gratefully acknowledge the Arkansas Children’s Infection Prevention and Control department for their contribution to data collection.

